# Improving prognosis of surrogate assay for breast cancer patients by absolute quantitation of Ki67 protein levels using Quantitative Dot Blot (QDB) method

**DOI:** 10.1101/2020.03.11.20034439

**Authors:** Junmei Hao, Yan Lv, Jiarui Zou, Yunyun Zhang, Shuishan Xie, Lili Jing, Fangrong Tang, Jiahong Lv, Xunting Wang, Jiandi Zhang

**Affiliations:** Dept. of Pathology, Yantai Affiliated Hospital of Binzhou Medical University, Yantai, P. R. China; Yantai Quanticision Diagnostics, Inc., a division of Quanticision Diagnostics, Inc. (US), Yantai, P. R. China; Dept. of Imaging, Linglong Yingcheng Hospital, Zhaoyuan, P.R. China

**Keywords:** Surrogate assay, adjusted surrogate assay, Ki67, QDB, IHC, quantitative

## Abstract

**Purpose:** The separation of Luminal A-like from Luminal B-like breast cancer subtypes in surrogate assay relies on Ki67 scores assessed by immunohistochemistry (IHC), a method known to be associated with subjectivity and inconsistency. We attempted to measure Ki67 levels absolutely, quantitatively and objectively in Formalin Fixed Paraffin Embedded (FFPE) specimens, and evaluate its influence on the performance of surrogate assay for breast cancer patients.

**Methods:** The Ki67 protein levels were assessed using both IHC and Quantitative Dot Blot (QDB) methods respectively in 253 specimens. These patients were assigned into Luminal A-like and Luminal B-like subtypes using either Ki67 score of 14% as cutoff in surrogate assay, or 2.31 nmole/g from QDB method as cutoff in adjusted surrogate assay. These two subtyping methods were compared with the Kaplan-Meier, univariate and multivariate survival analyses of the overall survival (OS) of Luminal-like patients.

**Results:** Ki67 levels measured using QDB method was highly correlated with those by IHC analysis (r=0.7, p<0.0001). The survival prediction for Luminal A-like patients was improved significantly in adjusted surrogate assay than surrogate assay (p=0.03 *vs* p<0.00052). The prediction of Hazard Ratio (HR) was also improve from 2.14 (95%CI: 0.89-5.11, p=0.087) to 6.89 (95%CI: 2.66-17.84, p<0.00001) in multivariate survival analysis.

**Conclusion:** Our study demonstrated that the inherent subjectivity and inconsistency associated with IHC analysis has adverse effect on the performance of surrogate assay.

This issue can be improved by objective and quantitative measurement of Ki67 levels with QDB method in daily clinical practice.

## Introduction

Microarray analysis of global gene expressions of breast cancer tissues leads to the identification the four intrinsic subtypes of luminal, Her2-like, basal-like and normal-like subtypes(1,2). This concept has been well accepted with several gene expression profiling (GEP)-based genetic tests developed to improve the prognosis and prediction of breast cancer patients in daily clinical practice(3).

Yet, GEP based genetic tests remains inaccessible to a lot of patients worldwide due to various reasons. As an alternative, surrogate assay, which is based on the protein expression levels of four biomarkers of Estrogen Receptor (ER), Progesterone Receptor (PR), Her2 and Ki67, has been used extensively in the world. Based on 2013 St. Gallen Consensus, the patients are categorized into Luminal A-like (LumA), Luminal B-like (LumB), Her2 positive (non-luminal), and Triple negative (ductal) subtypes. LumA patients, considered with best prognosis among all these subtypes, are defined as those patients with ER+, Her2-, PR > 20%, and Ki67 score < 14%. In some clinical practice, the cutoff of Ki67 score can also be 20%. LumB patients are comprised of Her2-(LumB_1_) and Her2+ (LumB_2_) sub-groups. LumB_1_ patients are ER+, Her2-, with Ki67 scores ≥14%, or PR<20%. LumB_2_ patients are both ER+ and Her2+, regardless of the Ki67 and PR statuses(4). Clearly, the Ki67 expression levels are critical to separate LumA from LumB_1_ in clinical practice.

Nonetheless, all these biomarkers are assessed using immunohistochemistry (IHC), a method known to be associated with subjectivity and inconsistency. In fact, among all four biomarkers used in surrogate assay, the standardization of Ki67 may be considered most difficult. Intensive efforts have been devoted to the standardization of this biomarker, yet, there are issues remains to be addressed(5–8). For example, it is still debatable if the hot spot should be included in the assessment(5,7). The intensity of the staining is also not considered when accessing the Ki67 levels using IHC method(8).

We hypothesized the inherent inaccuracy associated with Ki67 IHC scores may negatively affect the performance of surrogate assay. Therefore, we attempted to measure Ki67 levels absolutely and objectively using a recently developed Quantitative Dot Blot (QDB) method in Formalin Fixed Paraffin Embedded (FFPE) specimens(9–12). The specimens were separated into luminal A (LumA_*q*_) and luminal B subtypes (LumB_*q*_) based on the absolute quantitated Ki67 levels in these specimens in a retrospective study (Table 1). The prognosis of this adjusted surrogate assay is compared with that of surrogate assay to evaluate the potential impact of objective and absolute measurement of Ki67 on subtyping of Luminal-like patients in surrogate assay.

**Table 1:**
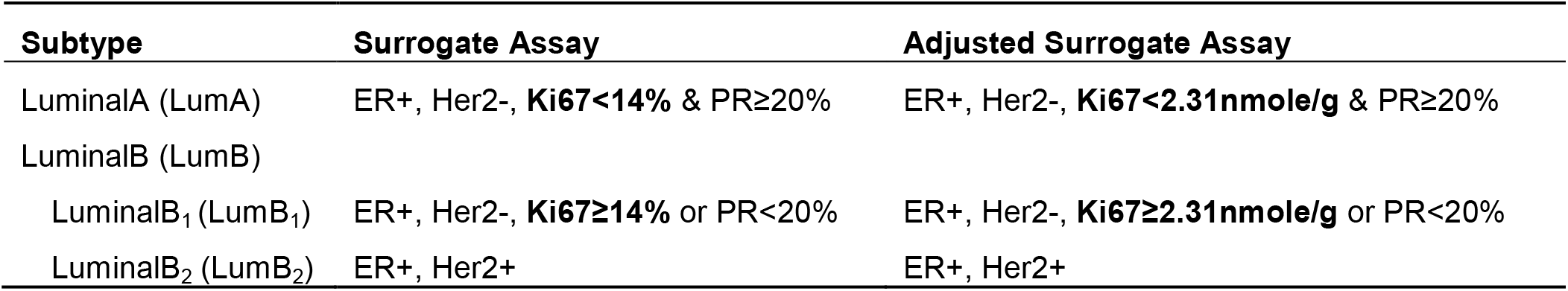
Subtyping of Luminal-like specimens.

## Materials and Methods

### Human subjects and human cell lines

A total of 253 Formalin Fixed Paraffin Embedded (FFPE) breast cancer tissue specimens were provided sequentially and non-selectively from 2010 to 2013 by Yantai Affiliated Hospital of Binzhou Medical University, Yantai, P. R. China. All the samples were obtained in accordance with the Declaration of Helsinki,and approved by the Medical Ethics Committee of Yantai Affiliated Hospital of Binzhou Medical University (Approval #: 20191127001). The informed Consent Forms were waived for archived specimens. The clinical information including the results of FISH analysis was obtained from medical records.

BT474 and 293T cell lysates were used as controls. Both of cell lines were purchased from the Cell Bank of Chinese Academy of Sciences (Shanghai, China), and maintained according to the provider’s instruction.

### General reagents

All general reagents used for cell culture were purchased from Thermo Fisher Scientific Inc. (Waltham, MA, USA), including the cell culture media and culture dishes. The protease inhibitors were purchased from Sigma Aldrich (St. Louis, MO, USA). All other chemicals were purchased from Sinopharm Chemicals (Beijing, P. R. China). QDB plates were manufactured by Quanticision Diagnostics, Inc. (RTP, USA). Mouse anti-Ki-67 antibody (clone MIB1) was purchased from ZSGB-BIO (Beijing, China). HRP-labeled Donkey Anti-Mouse IgG secondary antibody was purchased from Jackson Immunoresearch lab (Pike West Grove, PA, USA).

PCR reagents, restriction enzymes and T4 DNA ligase were purchased from Takara Inc. (Dalian, China). Competent cells E. coli DH5a and BL21(DE3) were from TransGen Biotech Inc. (Beijing, China). IPTG (Isopropyl β-D-1-thiogalactopyranoside) was purchased from Solarbio Inc. (Beijing, China). Nickel-His GraviTrap affinity column was purchased from GE Healthcare.

### Purification of recombinant Ki67 fragment

A DNA sequence corresponding to the 1162-1254AA of human MKI67 (NCBI #: NM_002417.4) was synthesized by Sangon Biotech. (Shanghai, China) and was inserted into PET32 (+) expression vector. The plasmid was verified by sequencing, and expressed in BL21 (DE3) competent cells. The cells were induced with IPTG, and total bacterial lysate was extracted in 10ml binding buffer (20 mM sodium phosphate, 500 mM NaCI, 20 mM imidazole, PH 7.4) before it was loaded onto a high affinity Ni2+ column pre-equilibrated with 10ml binding buffer. The recombinant protein was eluted with 3ml elution buffer (20 mM sodium phosphate, 500 mM NaCI, 250 mM imidazole, PH 7.4), and dialyzed in PBS (PH 7.4) at 4°C overnight. The purity of the protein was examined by a 12% SDS-PAGE gel at 80%, and the purified protein was stored at - 80°C in small aliquot with 20% glycerol.

### Preparation of FFPE and cell lysates

To extract total protein, 2×5 μm FFPE slices were first de-paraffinized, then solubilized with lysis buffer (50 mM HEPES, 137 mM NaCl, 5 mM EDTA, 1 mM MgCl, 10 mM Na_2_P_2_O_7_, 1% TritonX-100, 10% glycerol). Total protein concentration was measured using Pierce BCA protein assay kit in accordance to the manufacturer’s instructions. BT474 and 293T cells were fixed in Formalin Solution for 30 mins before they were lysed in the same lysis buffer with protease inhibitors. The supernatants were collected after centrifugation and the total amount of proteins was measured using BCA protein assay kit by following manufacturer’s instructions.

### QDB analysis

FFPE tissue lysates were adjusted to 0.5 μg/μl according to the BCA assay. Sample pool was prepared by mixing tissue lysates from four FFPE tissue specimens with IHC score above 70%, and was serially diluted side by side with the recombinant Ki67 protein for defining the sample and standard curve of QDB analysis.

The QDB process was described in detail elsewhere with slightly modifications(9,10). In short, the final concentration of the FFPE tissue lysates was adjusted to 0.25 μg/μl, and 2 μl/unit was used for QDB analysis as well as a serially diluted recombinant protein in triplicate. The QDB plate was then dried for 1 hour at RT, soaked in 20% methanol for 10s, rinsed once with TBST, and then blocked in 4% non-fat milk for an hour. Anti-Ki67 antibody was diluted at 1:1000 in blocking buffer, and incubated with QDB plate at 100 μl/well for overnight at 4°C. Afterward, the plate was rinsed twice with TBST and washed 3X10 mins and a donkey anti-mouse secondary antibody was incubated with the plate for 4 hours at RT. The plates were briefly rinsed twice with TBST, and washed 5X10mins. Finally, the QDB plate was inserted into a white 96-well plate pre-filled with 100 μl/well ECL working solution for 3 mins. The chemiluminescence signals of the combined plate were quantified by using the Tecan Infiniti 200pro Microplate reader with the option “plate with cover”.

The consistency of the experiments was ensured by including BT474 and 293T cell lyastes of known Ki67 levels in all the experiments. The result was considered valid when the calculated Ki67 level of BT474 and 293T was within 20% of known Ki67 level. The absolute Ki67 level was determined based on the dose curve of protein standard. Ki67 level less than 25 pg (about 1.4 nmole/g) was defined as non-detectable, and entered as 0 for data analysis.

#### IHC analysis

Immunohistochemistry for ER, PR, HER2, and Ki67 was performed concurrently on serial sections with the standard streptavidin–biotin complex method with 3, 3′-diaminobenzidine as the chromogen. Staining for ER, PR, and HER2 interpretation was performed by following the Dako autostainer link 48 manual (Ft. Collins, Colorado, CO). ER antibody (clone SP1) and PR antibody (clone SP2) from MXB Biotechnologies, HER2 antibody (polyclonal A0485) from Dako, and Ki67 (Clone SP6) from LBP (www.gzlbp.com) were all at 1:100 dilution after antigen retrieval in 0.05 M Tris buffer (pH 9.0) with heating to 95°C for 20 minutes. Biomarker expressions from immunohistochemistry assays were scored by three pathologists (JM Hao, JR Zou & LL Jing), who were blinded to the clinicopathological characteristics and outcomes and who used previously established/published criteria for biomarker expression levels routinely used in daily clinical practice. Tumors were considered positive for ER if immunostaining was observed in more than 1% of tumor nuclei, as recommended by ASCO/CAP guidance. Tumors were considered positive for HER2 if immunostaining was scored as 3+ according to HercepTest criteria or FISH test positive. Ki67 and PR were visually scored for percentage of tumor cell nuclei with positive immunostaining above the background level.

### Statistical analysis

For Ki67 measured with QDB method, Graph Pad 7 software (La Jolla, CA, USA) was used for data analysis in this study, and the results were presented as Mean ± SEM. The correlation between IHC score and Ki67 levels from QDB analysis was analyzed using Pearson’s correlation coefficient analysis.

The survival analyses for this study were done using R version 3.6.2. The end-point was overall survival defined as the time between breast cancer surgery and death or last follow-up. Patients still alive at last study follow-up (April 1st, 2019) were censored. 12 patients lost to follow up in Luminal group were treated as missing data, and was not included in the corresponding analyses (supplemental table 1). The strength of the agreement among K67 IHC scores from three pathologists was assessed by Fleiss’s Kappa analysis.

The Ki67 levels measured by QDB method and IHC method were dichotomized for OS by using optimal cutoff values determined by the “surv_cutpoint” function of the “surviminer” R package respectively. For Ki67 levels measured with QDB method, a cutoff at 2.31 nmole/g was obtained to distinguish luminal-like breast cancers into Luminal A-like and Luminal B_1_-like subtypes. The same analysis was also performed using average Ki67 scores from three pathologists, and an optimized cutoff of 2.67% were obtained. However, the well-accepted 14% cutoff was used primary in this study unless specified in the text. All the Overall survival analyses were visualized by Kaplan-Meier method, and comparisons were performed by log-rank test.

Univariate Cox proportional hazard models fitted overall survival were employed for hazard ratio (HR) and corresponding 95% confidence intervals (CIs) estimation. Multivariable Cox models were utilized to examine the association between subtypes and OS, adjusting for other clinical variables, such as age, node status, tumor size, tumor grade, and type of treatment. Residuals that are analogous to the Schoenfeld residuals in Cox models were used to check the proportionality assumption. P values of less than .05 were considered statistically significant

## Results

### Measurement of Ki67 levels with QDB method

A QDB-based high throughput immunoassay for absolute quantitation of Ki67 levels in FFPE specimens was developed first by defining the linear range of total tissue lysates and recombinant Ki67 protein standards using a clinically validated antibody, MIB1 (supplemental figure 1). Total tissue lysates from 4 FFPE specimens with Ki67 score>90% were pooled together, and used to define the linear range of the assay.

**Fig. 1:**
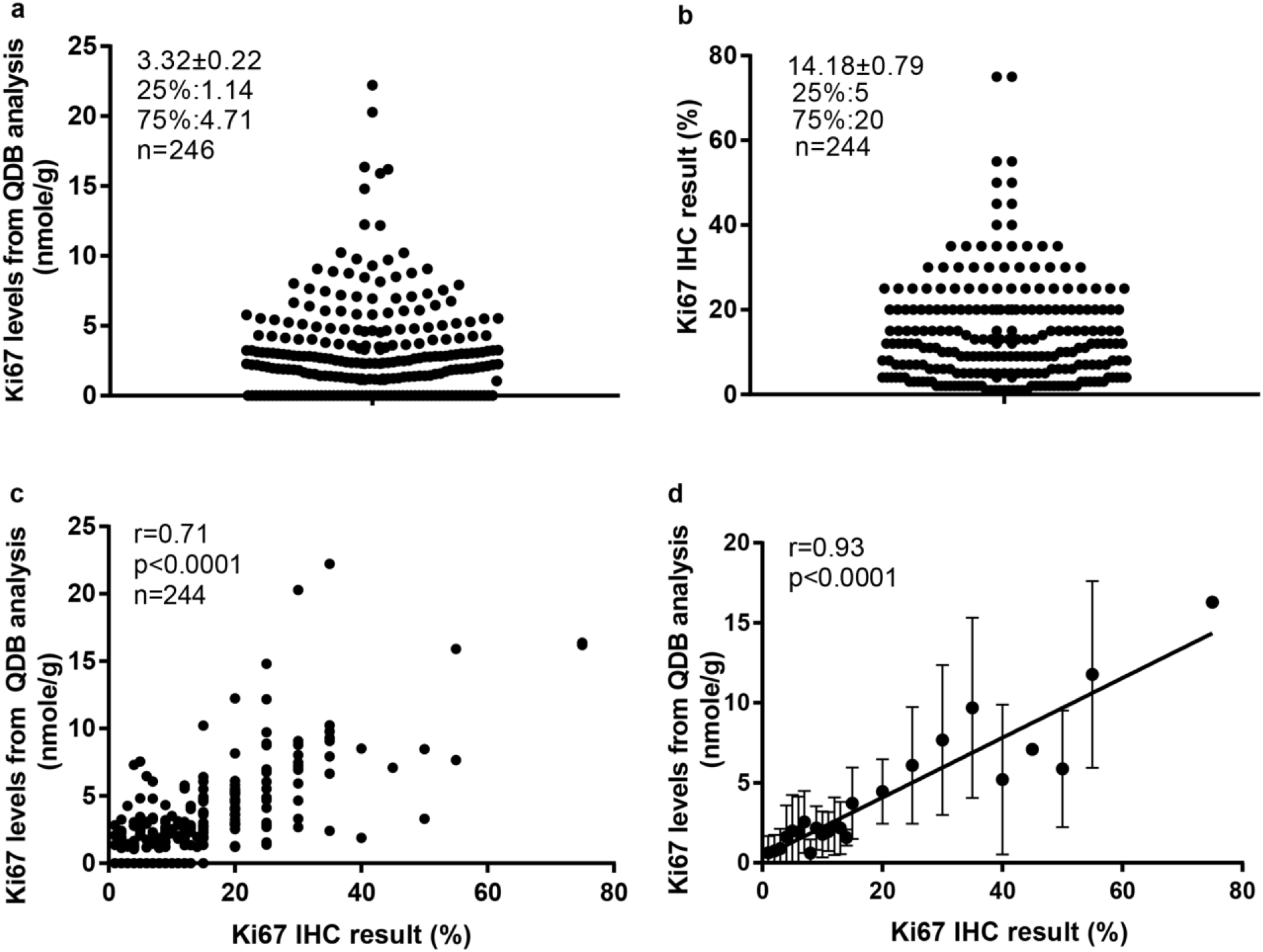
Measuring Ki67 levels absolutely and quantitatively in 253 FFPE specimens using QDB method and their correlations with Ki67 scores from IHC analysis. Total lysate was extracted from 2X5 μm FFPE slices individually, and 0.5 μg/specimen was used for QDB measurement using Mouse anti-Human Ki67 monoclonal antibody (MIB1). These specimens were also assessed with IHC analysis, with each IHC stained slides assessed by three pathologists independently. The Ki67 scores used in the study were averages of three assessments. (a) Distribution of quantitatively measured Ki67 levels among these specimens. (b) Distribution of Ki67 scores from IHC analysis among 244 specimens. (c) Correlation analysis of the results from QDB and IHC analyses using Pearson’s correlation analysis with r=0.72, p<0.0001; (d) These specimens were sub-grouped based on their respective Ki67 scores. The subgroup averages of the Ki67 levels from QDB measurements were used for correlation analysis with Ki67 scores from IHC analysis using Pearson correlation analysis with r=0.93, p<0.0001.

All 253 FFPE specimens provided were measured using QDB method, and Ki67 levels were found to distribute between 0 (undetectable level) to 22.21 nmole/g, with average at 3.32±0.22 nmole/g. One fifth (57 out of 246) of the samples were found with Ki67 levels below detection level (Fig. 1a).

Among 253 specimens provided, 244 were provide with Ki67 scores from three pathologists independently assessing the same set of IHC stained slides in the local hospital. The average of the Ki67 scores of these specimens were used in this study. We found the highest IHC score was at 75%, and the lowest at 1%, with average at 14.17±0.79% (Fig. 1b). Correlation analysis was performed using results from QDB and IHC methods, and we obtained r=0.72, p<0.0001 using Pearson’s correlation analysis (Fig 1c). In an attempt to reduce the potential interference from the subjectivity inherently associated with IHC analysis, we also sub-grouped these specimens by their IHC scores. As expected, the correlation between the subgroup averages of the absolute Ki67 levels from QDB method with the matching IHC scores was increased to r=0.93, p<0.00001 when analyzed using Pearson’s correlation analysis (Fig. 1d).

The 244 Specimens provided with Ki67 scores from IHC analysis were also accompanied by IHC results for ER, PR, and Her2. For specimens with Her2 score of 2+, results from FISH analysis were used to differentiate Her2+ from Her2-specimen. Therefore, we were able to assign these 244 specimens into 155 luminal-like subtype, 31 HER2-like subtype, and 53 Triple Negative subtype based on 2013 St. Gallen consensus(4). The remaining 5 specimens cannot be subtyped based on this consensus (Fig. 2). When using Ki67 score at 14% as cutoff, we further separated the luminal-like subtype into 66 Luminal A-like subtype and 89 luminal B-like subtype. When 20% Ki67 score was used as cutoff, the number was changed to 76 Luminal A-like subtype, and 79 Luminal B-like subtype.

**Fig. 2:**
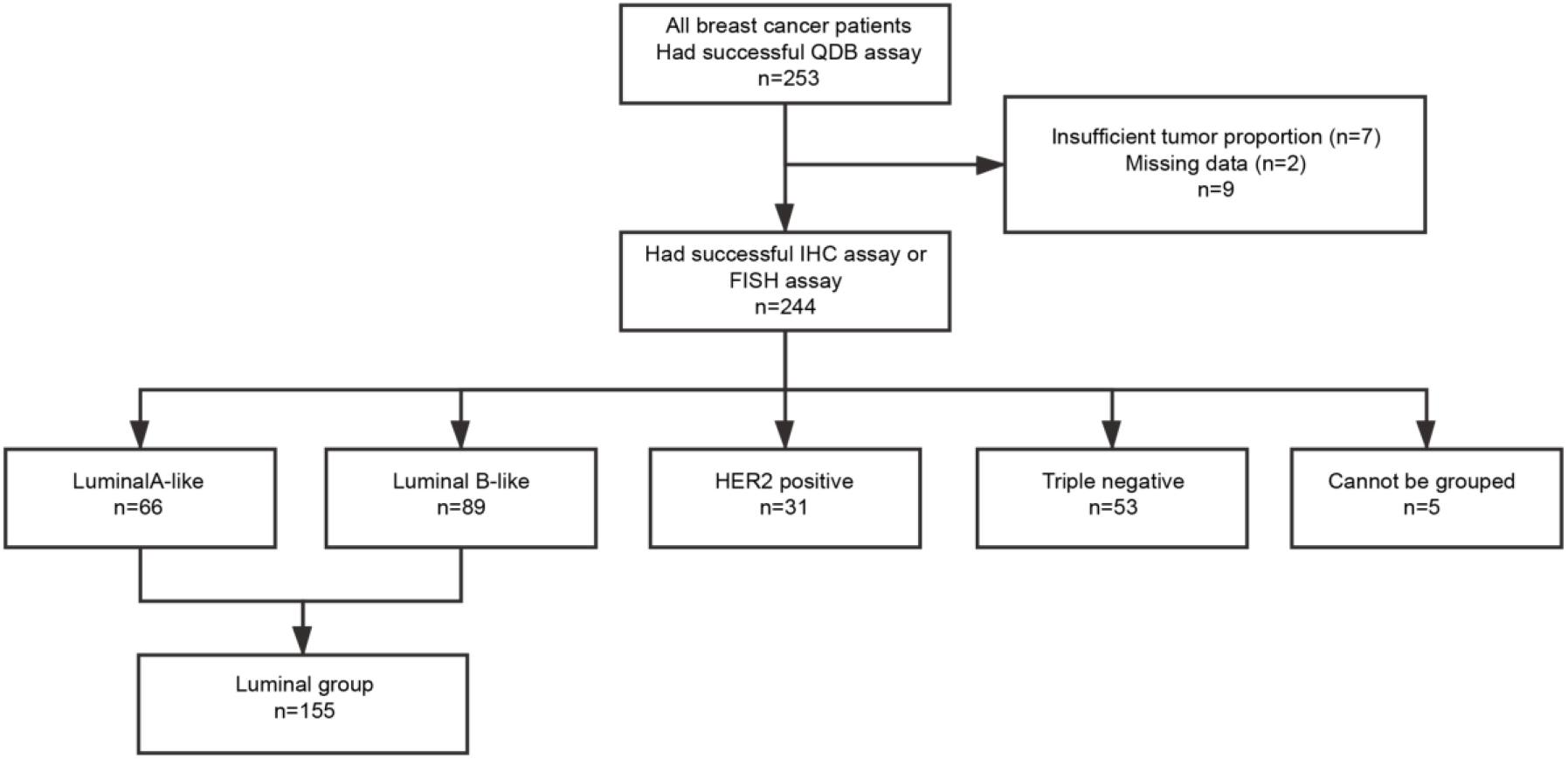
Flow diagram of patient selections for the study.

The clinicopathological parameters of the 155 luminal-like specimens were listed in table 2. In this retrospective study, no recurrence data were available for further analysis. Therefore, we focused our study on the Overall Survival (OS) of the patients only. In addition, among the patients with known treatments, majority of them received chemotherapy with or without endocrine therapy. Only two patients received endocrine therapy alone.

**Table 2:** Clinicopathological characteristics of 155 specimens with Luminal-like breast cancer subtypes.

| Characteristics | Number of Cases (%) |
| --- | --- |
| **Age** |  |
| <50 | 69(44.52) |
| ≥50 | 86(55.48) |
| **Treatment** |  |
| Endo | 2(1.29) |
| Chemo | 90(58.06) |
| Endo&Chemo | 41(26.45) |
| Unknown | 22(14.19) |
| **Node Status** |  |
| N0 | 75(48.39) |
| N1 | 56(36.13) |
| N2 | 10(6.45) |
| N3 | 8(5.16) |
| Unknown | 6(3.87) |
| **Tumor Size** |  |
| T1 | 54(34.84) |
| T2 | 91(58.71) |
| T3 | 7(4.52) |
| Unknown | 3(1.94) |
| **Grade** |  |
| II | 83(53.55) |
| III | 52(33.55) |
| Unknown | 20(12.9) |
| **Surrogate Assay (14% Ki67 cutoff)** |  |
| LumA<sub>i</sub> | 66(42.58) |
| LumB<sub>i</sub> | 89(57.42) |
| LumB<sub>1i</sub> | 70(45.16) |
| LumB<sub>2i</sub> | 19(12.26) |
| **Surrogate Assay (20% Ki67 cutoff)** |  |
| LumA<sub>i</sub> | 76(49.03) |
| LumB<sub>i</sub> | 79(50.97) |
| LumB<sub>1i</sub> | 60(38.71) |
| LumB<sub>2i</sub> | 19(12.26) |

To evaluate the influence of objectively quantitated Ki67 levels on the prognostic effect of surrogate assay, we replaced the Ki67 scores used in surrogate assay to distinguish luminal A-like subtype from luminal B-like subtype by an optimized absolute Ki67 level at 2.31 nmole/g, and named this revised method the adjusted surrogate assay (table 1). The 2.31 nmole/g cutoff used in adjusted surrogate assay was obtained through calculation using the “surv_cutpoint” function of the “suvminer” R package in combination with the Overall Survival (OS) of these patients.

The same calculation was also performed with surrogate assay to obtain the optimum cutoff for Ki67 score from IHC analysis, and we obtained the optimum cutoff at 2.67%. At this value, the 10 year survival probability was at 96% for Luminal A-like subtype and 72% for Luminal-B subtype, with p=0.021(data not shown). However, only 26 specimens were assigned to Luminal A-like subtype, in comparison to 61 when 14% was used as cutoff. Therefore, this 2.67% was not considered a valid cutoff in this study.

As shown in Fig. 3, using 2.31 nmole/g as cutoff, the luminal A-like subtype from adjusted surrogate assay (LumA_*q*_) has 91% survival probability over 10 years, while LumB_*q*_ subtype has 63% survival probability over the same period. The p value from Log rank test for adjusted surrogate assay was 0.00052. On the other hand, using Ki67 score of 14% as cutoff, the luminal A-like subtype in surrogate assay (LumA_*i*_) has 10 year survival probability of 88%, while LumB_*i*_ has 68% survival probability over the same period. The p value from log rank test for surrogate assay was 0.031. When Ki67 score of 20% was used as cutoff, the 10 year survival probability for LumA_*i*_ was 84%, in contrast to 70% for LumB_*i*_, with the p value from Log rank test at 0.10 (supplemental figure 2). Therefore, in the following studies, the 2.31 nmole/g value was used as cutoff in adjusted surrogate assay while the 14% Ki67 was used as the cutoff in Surrogate assay.

**Fig. 3:**
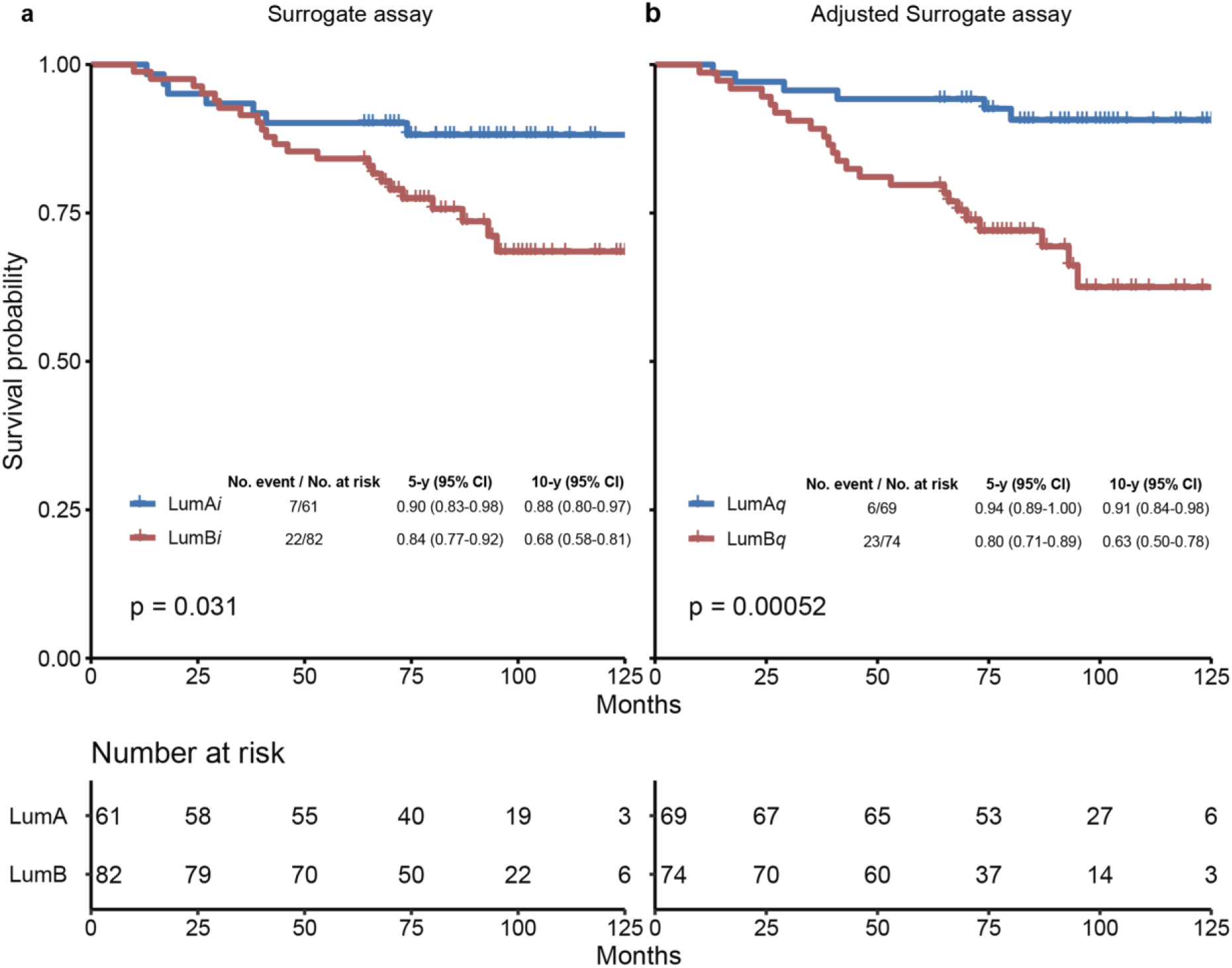
Kaplan-Meier curves for overall survival according to breast cancer subtypes assigned by surrogate assay (a) or adjusted surrogate assay (b) in Luminal-like subtypes. The Ki67 score of 14% was used as cutoff in surrogate assay based on recommendations from 2013 St. Gallen Consensus (a). The Ki67 level of 2.31 nmole/g was used as cutoff determined by the “surv_cutpoint” function of the “surviminer” R package in adjusted surrogate assay (b). The 5 year and 10 year survival probabilities, and the p values from Log Rank test were provided for both Surrogate assay and Adjusted Surrogate assay respectively. LumA, Luminal A-like subtype; LumB, Luminal B-like subtype; LumA_*i*_ and LumB_*i*_, Luminal A-like and B-like subtypes by surrogate assay; LumA_*q*_ and LumB_*q*_, Luminal A-like and B-like subtypes by adjusted surrogate assay; CI, confidence interval.

The surrogate assay was compared next with adjusted surrogate assay in univariate cox regression analysis, and we found that adjusted surrogate assay provided improved prognosis for Luminal-like breast cancers with HR at 4.39 (95%CI, 1.78-10.81, p=0.0013) than that of surrogate assay with HR at 2.46 (95%CI, 1.05-5.75, 0.0385) (Table 3).

**Table 3:** Univariate cox regression analysis of Overall Survival (OS) of Luminal-like patients subtyped by surrogate assay or adjusted surrogate assay respectively. The 155 Luminal-like patients were subtyped based on Ki67 scores from IHC analysis (surrogate assay) or absolute Ki67 levels from QDB analysis (adjusted surrogate assay) respectively, and univariate cox regression analysis for OS was performed for these two subtyping methods respectively.

| Variable | HR | 95%CI | P-value |
| --- | --- | --- | --- |
| Surrogate Assay | 2.46 | 1.05-5.75 | 0.0385 |
| Adjusted Surrogate Assay | 4.39 | 1.78-10.81 | 0.0013 |

The prognostic values of both surrogate assay and adjusted surrogate assay were also investigated respectively in the multivariate cox regression analysis to include age, treatment, node status, tumor size, tumor grade in the analysis. We found the HR of surrogate assay at 2.14 (95%CI, 0.89-5.11, p=0.0873) while that of adjusted surrogate assay was at 6.89 (95%CI, 2.66-17.84, p=0.0001) (Table 4). In addition, in both analyses, age and node status were found to be an independent prognostic factor.

**Table 4:**
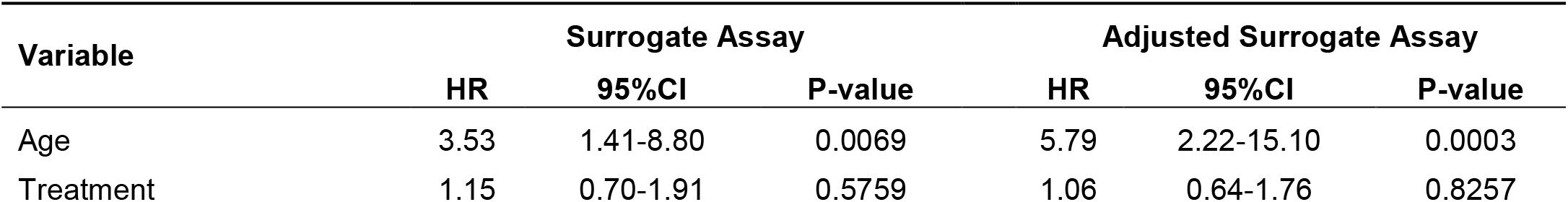
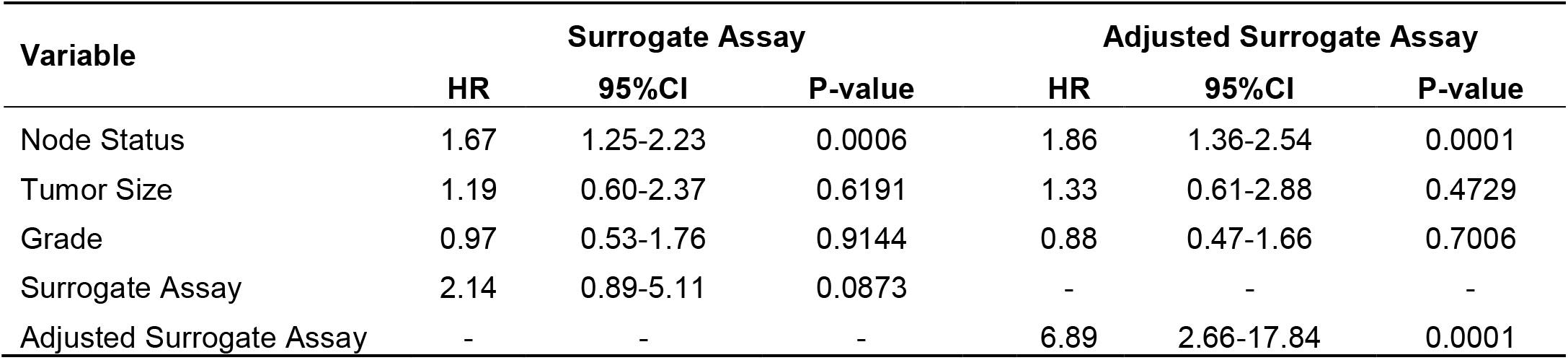
Multivariate cox regression analysis of OS of Luminal-like patients subtyped by surrogate assay or adjusted surrogate assay respectively. The 155 Luminal-like patients were subtyped based on surrogate or adjusted surrogate assay respectively, and analyzed together with age, treatment, node status, tumor size, and histological grade in multivariate cox regression analysis for OS of these patients.

| Variable | Surrogate Assay |  |  | Adjusted Surrogate Assay |  |  |
| --- | --- | --- | --- | --- | --- | --- |
|  | HR | 95%CI | P-value | HR | 95%CI | P-value |
| Age | 3.53 | 1.41-8.80 | 0.0069 | 5.79 | 2.22-15.10 | 0.0003 |
| Treatment | 1.15 | 0.70-1.91 | 0.5759 | 1.06 | 0.64-1.76 | 0.8257 |
|  | HR | 95%CI | P-value | HR | 95%CI | P-value |
| Node Status | 1.67 | 1.25-2.23 | 0.0006 | 1.86 | 1.36-2.54 | 0.0001 |
| Tumor Size | 1.19 | 0.60-2.37 | 0.6191 | 1.33 | 0.61-2.88 | 0.4729 |
| Grade | 0.97 | 0.53-1.76 | 0.9144 | 0.88 | 0.47-1.66 | 0.7006 |
| Surrogate Assay | 2.14 | 0.89-5.11 | 0.0873 | - | - | - |
| Adjusted Surrogate Assay | - | - | - | 6.89 | 2.66-17.84 | 0.0001 |

Next, we tried to understand what caused this difference by comparing the luminal A-like and Luminal B-like subtypes from surrogate assay (LumA_*i*_ and LumB_*i*_) with those from adjusted surrogate assay (LumA_*q*_ and LumB_*q*_) in Table 5. The specimens assigned to luminal A-like subtype by both assays were named A_i_A_q_. and specimens assigned to Luminal B-like subtypes by both assays as B_i_B_q_. Likewise, those assigned by surrogate assay to A-like subtype, but not by adjusted surrogate assay was named as A_i_B_q_, and those assigned by adjusted surrogate assay to Luminal-A like subtype, but not by surrogate assay were named B_i_A_q_. We found more specimens were assigned to Luminal A-like subtype by adjusted surrogate assay than surrogate assay (76 *vs* 66). The overall concordance rate between surrogate assay and adjusted surrogate assay was 75.5%.

**Table 5:**
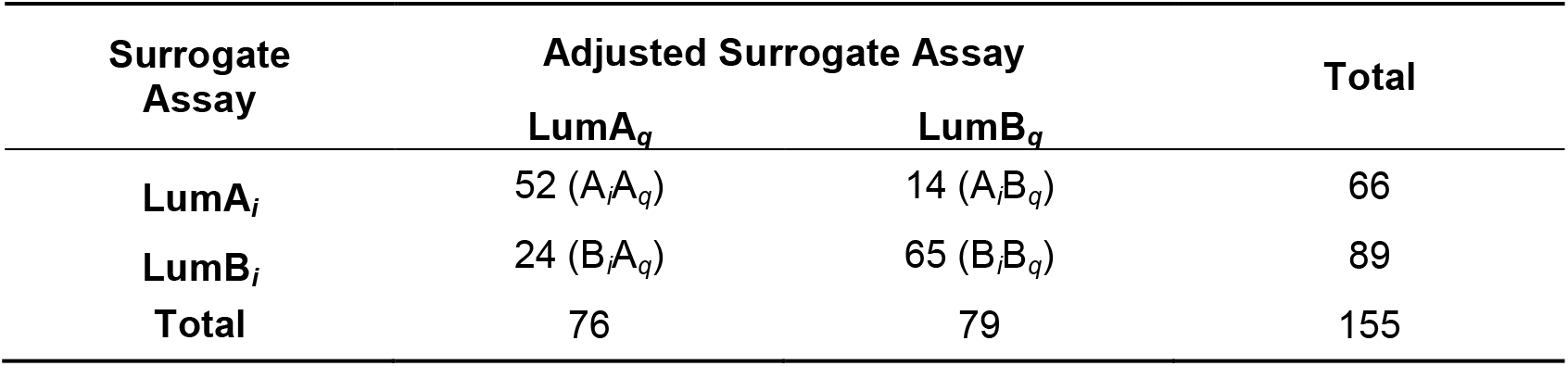
Concordance of surrogate assay and adjusted surrogate assay. The 155 Luminal-like patients were subtyped into 66 Luminal A-like (LumA_*i*_) and 89 Luminal B-like (LumB_*i*_) subtypes based on surrogate assay, or 76 Luminal A-like (LumA_*q*_) and 79 Luminal B-like (LumB_*q*_) subtypes based on adjusted surrogate assay. There were 52 patients assigned by both surrogate assay and adjusted surrogate assay as Luminal A-like subtype (A_i_A_q_), 65 assigned by both surrogate assay and adjusted surrogate assay as Luminal B-like subtype (B_i_B_q_), 14 assigned by surrogate assay as Luminal A-like subtype, but assigned as Luminal B-like subtype by adjusted surrogate assay (A_i_B_q_), and 24 assigned by surrogate assay as Luminal B-like subtype, but as 540 Luminal A-like subtype by adjusted surrogate assay (B_i_A_q_). The overall concordance of 541 these two methods were (52+65)/155=75.5%.

| Surrogate Assay | Adjusted Surrogate Assay |  | Total |
| --- | --- | --- | --- |
|  | LumA<sub>q</sub> | LumB<sub>q</sub> |  |
| LumA<sub>i</sub> | 52 (A<sub>i</sub>A<sub>q</sub>) | 14 (A<sub>i</sub>B<sub>q</sub>) | 66 |
| LumB<sub>i</sub> | 24 (B<sub>i</sub>A<sub>q</sub>) | 65 (B<sub>i</sub>B<sub>q</sub>) | 89 |
| Total | 76 | 79 | 155 |

In Fig. 4, we performed the survival analyses of these four subgroups using Kaplan-Meier survival analysis, and found the subgroup assigned by both methods to Luminal B-like subtype had the worst prognosis with the 10 year survival probability at 59% (B_i_B_q_). The subgroup assigned by both methods to Luminal A-like subtype had the best 10 year survival probability of 91%. In addition, the 10 year survival probability of B_i_A_q_, the subgroup assigned to Luminal A-like subtype only by adjusted surrogate assay, was very close to that of A_i_A_q,,_ the subgroup assigned by both assays to Luminal A-like subtype, at 90%.

**Fig. 4:**
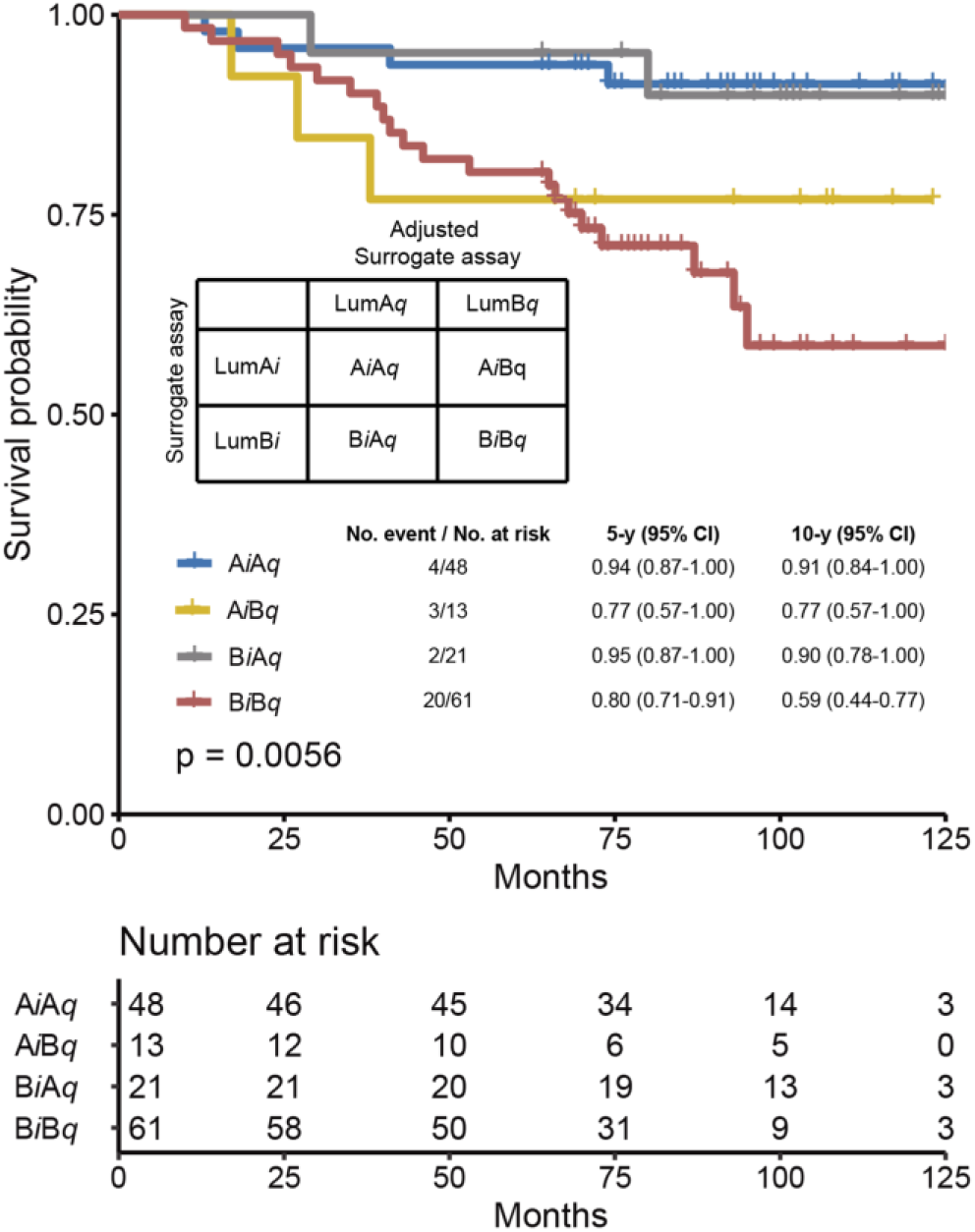
Further dissecting the differences between surrogate assay and adjusted surrogate assay. The specimens were further sub-grouped into A_i_A_q_ and B_i_B_q_ subgroups, representing specimens assigned as Luminal A-like subtype and Luminal B-like subtype by both assays; A_i_B_q_, representing specimens assigned as Luminal A subtype by surrogate assay, but as Luminal B subtype by adjusted surrogate assay; and B_i_A_q_, representing specimens assigned as Luminal B-like subtype by surrogate assay, but as Luminal A-like subtype by adjusted surrogate assay. The Overall survival analysis was performed with these four subgroups using Kaplan-Meier survival analysis, with survival probability for each individual subgroup provided in the figure. The p value was calculated with Log Rank test.

We also separate the patients by treatments to minimize the influence of treatment on the survival probability of each subtype (Fig. 5). As mentioned previously, these patients were treated predominantly with chemotherapy. Among 85 patients, 34 were assigned to Luminal A-like subtype by surrogate assay in comparison to 41 by adjusted surrogate assay. Consistent with the overall performance, adjusted surrogate assay presented significantly better prognosis than surrogate assay, with the 10 year survival probability at 100% for LumA_*q*_ *vs* 53% for LumB_*q*_, p<0.0001. In contrast, Luminal A-like subtypes for surrogate assay (LumA_*i*_) had the 10 year survival probability at 94 *vs* 69% for LumB_*i*_, p=0.037 (Fig. 5a and 5b).

**Fig 5:**
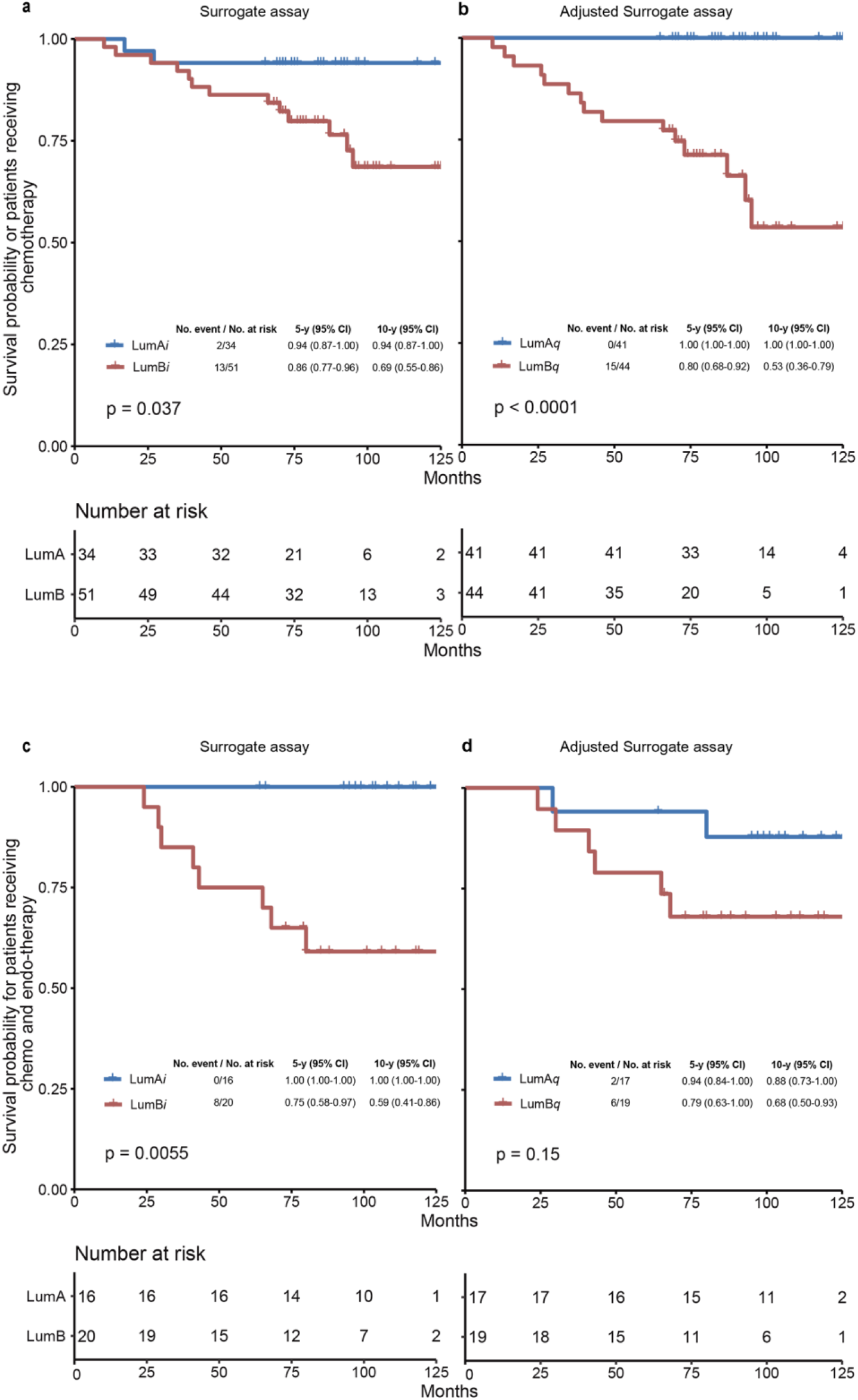
Comparison of surrogate assay and adjusted surrogate assay by treatments. The FFPE specimens were separated into groups receiving chemotherapy alone (a & b) and those receiving combined chemo and endotherapies (c & d) respectively. Within each group, these specimens were further subtyped into Luminal A-like and Luminal B-like subtypes using surrogate assay (a & c) and adjusted surrogate assay (b & d) respectively. The Kaplan-Meier survival analyses were performed for these four subtyping groups respectively, with 5 and 10 year survival probabilities provided in the figure. The p values were calculated using Log Rank test. LumA, Luminal A-like subtype; LumB, Luminal B-like subtype; LumA_*i*_ and LumB_*i*_, Luminal A-like and B-like subtypes by surrogate assay; LumA_*q*_ and LumB_*q*_, Luminal A-like and B-like subtypes by adjusted surrogate assay; CI, confidence interval; P, log-rank p-value.

However, for the 35 patients receiving both chemotherapy and endotherapy treatment, adjusted surrogate assay showed no advantage over surrogate assay. In fact, adjusted surrogate assay was unable to separate patients effectively, while surrogate assay offers significant advantage (the 10 year survival probability for LumA_*q*_ were 88% *vs* LumB_*q*_ at 68%, p=0.15, in comparison to 100% *vs* 59% for LumA_*i*_ *vs* LumB_*i*_, p=0.0055 (Fig. 5c and 5d).

The potential influence of node status was also investigated in this study by dividing patients into N0 group (no positive lymph node) and N1 (patients with 1 to 3 positive lymph nodes), and analyzed their 10 year survival probability using Kaplan-Meier analysis. Again, adjusted surrogate assay showed better prognosis than surrogate assay in both cases, with 10 year survival probability of 97% *vs* 72% for LumA_q_ *vs* LumB_q_, p=0.023 for adjusted surrogate assay, in comparison for 93% *vs* 80% for LumA_i_ *vs* LumB_i_, p=0.31 for surrogate assay for N0 patients; and 90% *vs* 63% LumA_q_ *vs* LumB_q_, p=0.026 for adjusted surrogate assay in comparison to 90% *vs* 66% for LumA_i_ *vs* LumB_i_, p=0.1 for surrogate assay for N1 patients. The patient numbers for N2 and N3 statuses were insufficient for further analysis (Fig. 6).

**Fig. 6:**
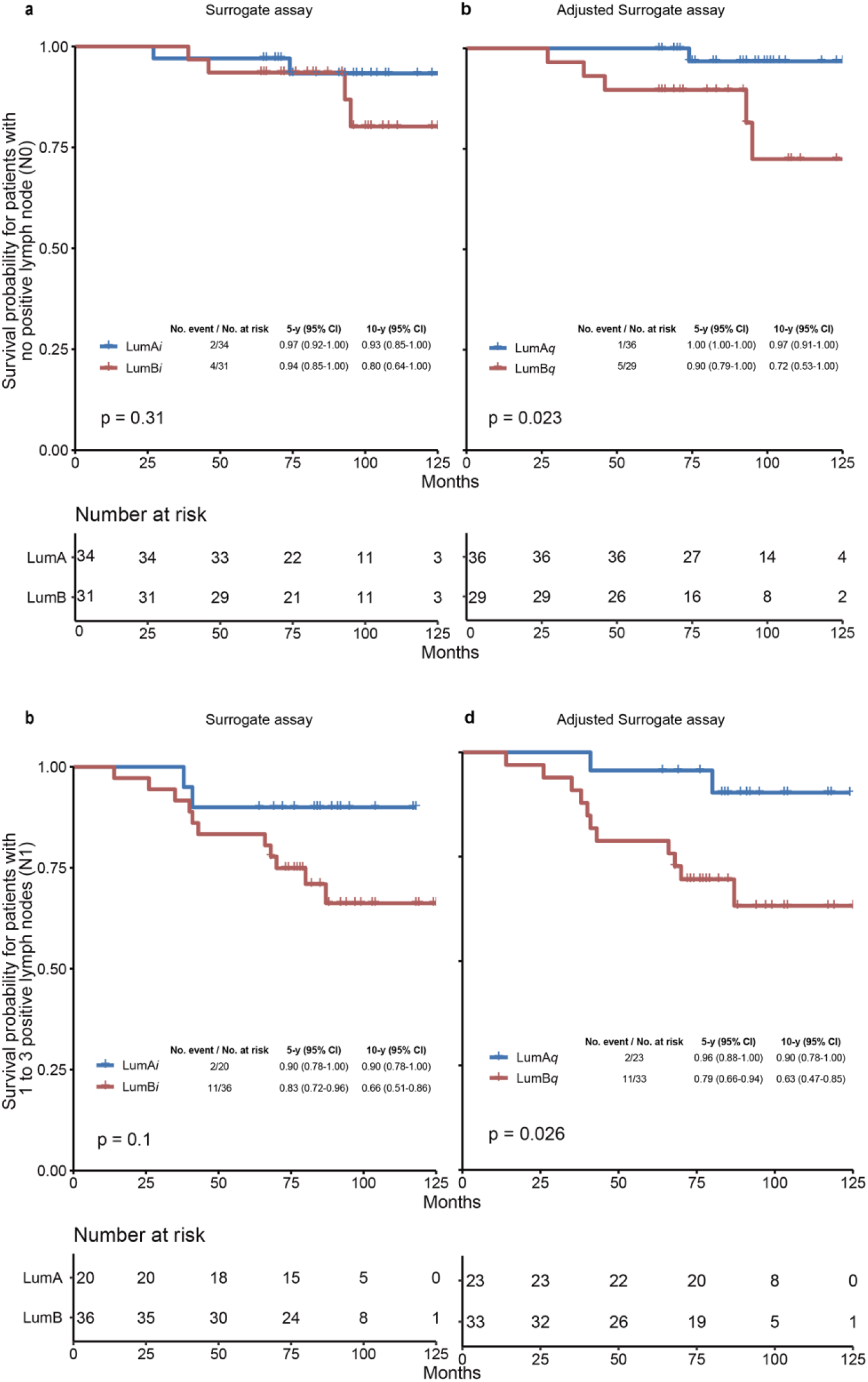
Comparison of surrogate assay and adjusted surrogate assay by node statuses. The FFPE specimens were grouped into N0 groups (patients with no positive lymph node) and N1 (patients with 1 to 3 positive lymph nodes) respectively. Within each group, these specimens were further subtyped into Luminal A-like and Luminal B-like subtypes using surrogate assay (a & c) and adjusted surrogate assay (b & d) respectively. The Kaplan-Meier survival analyses were performed for these four subtyping groups respectively, with 5 and 10 year survival probabilities provided in the figure. The p values were calculated using Log Rank test. LumA, Luminal A-like subtype; LumB, Luminal B-like subtype; LumA_*i*_ and LumB_*i*_, Luminal A-like and B-like subtypes by surrogate assay; LumA_*q*_ and LumB_*q*_, Luminal A-like and B-like subtypes by adjusted surrogate assay; CI, confidence interval; P, log-rank p-value.

## Discussion

In this study, by using objectively quantitated Ki67 levels to replace Ki67 score in surrogate assay, we showed that inherent subjectivity and inconsistency of IHC analysis impaired the performance of surrogate assay in identifying luminal A breast cancer subtype from Luminal B-subtype. Thus, we propose revise the surrogate assay to use quantitative Ki67 levels in separating Luminal A-like from Luminal B-like breast cancer subtype. We believe this modification should significantly improve the accuracy and consistency of surrogate assay in daily clinical practice worldwide.

Indeed, the standardization, or lack of standardization of Ki67 in clinical practice, is a challenge facing the whole medical community. Yet, until now, no significant progress has been made so far. Our study suggests that the issue may be at the methodology. The significantly improved prognosis in adjusted surrogate assay suggests strongly that high throughput immunoassay should be the right choice for Ki67 standardization in daily clinical practice.

Indeed, one of the biggest challenge for IHC analysis is tumor heterogeneity, a phenomenon commonly observed in solid tumor. In IHC analysis, a typical field of view is a circle of 0.5mm in radius. In other word, it is only 1/400 of a typical FFPE slice of 1cmX1cm. It is hard to imagine that by random selection of three field of views, as recommended by International Ki67 in Breast Cancer Working Group(5), the overall Ki67 levels of the FFPE specimens would be reflected faithfully. In contrast, the whole tissue is homogenized in QDB method to reduce the influence of tumor heterogeneity to the minimum. Needless to say, this is at the cost of its morphological features. Nonetheless, results from our studies strongly suggest that the overall Ki67 levels from QDB method were more relevant to the performance of surrogate assay than the observational results from IHC analysis.

The performance of surrogate assay is also seriously affected by the subjectivity of the assay. In this study, we managed to reduce its influence by requested three pathologists to judge the Ki67 scores in these samples independently, and use the average Ki67 scores in our analyses throughout the study. However, the personal biases are clearly noticeable. The IHC results from these three pathologists were analyzed with Fleiss Kappa correlation analysis, and we had κ=0.633 among these pathologists. We also assigned the patients into Luminal A-like and Luminal B-like subtypes using the IHC results from these three pathologists respectively, and found the p values from log rank tests were at 0.2, 0.018 and 0.1 between Luminal A-like and Luminal B-like subtypes in Kaplan-Meier survival analysis of these patients.

Perceivably, by including more pathologists in the analysis, the subjectivity of IHC analysis should be minimized. This assumption may find its support in Fig. 1d, where we showed significantly increased correlation between QDB and IHC when the subgroup averages of Ki67 levels from QDB method were used in our correlation analysis. Nonetheless, even using Ki67 scores averaged from three pathologists, the surrogate assay still performed poorly than adjusted surrogate assay in this study.

Admittedly, there are several obvious limitations with this study. As a retrospective study, we were unable to evaluate the performance of surrogate assay and adjusted surrogate assay on the prognosis of the recurrence of the disease for lacking of relevant data. Our conclusions are also affected by the small sample size in this study. It remains questionable if this conclusion can be held up with more FFPE specimens in the study. Future studies are needed to validate our conclusion in the near future.

It is also questionable if our proposed 2.31 nmole/μg cutoff remains effective in a larger cohort study. However, we are optimistic in this regard. We have demonstrated that the Ki67 levels measured with QDB method, when averaged by the Ki67 scores, are tightly associated with the results from IHC analysis. We speculate that this is because by including IHC scores from multiple pathologists at multiple occasions, the subjectivity of this method is maximally canceled out with each other. In this regard, the 14% cutoff in the surrogate assay was developed from a large population study. Thus, it should be the optimized value to achieve best prognosis for surrogate assay. Interestingly, in our correlation studies of QDB method and IHC methods with more than 2000 FFPE specimens analyzed, we found the 2.31 nmole/g matched to the subgroup average of Ki67 levels around 10% ∼ 15% Ki67 scores (unpublished data). Therefore, we expected this value to be in close proximity to the optimized cutoff in the future.

We also observed that adjusted surrogate assay failed to show any advantage in separating Luminal A-like patients from Luminal B-like patients among patients receiving both endo and chemotherapies. We do not have a good explanation to this inconsistency at this point. However, this observation suffered from very small sample size (n=35). The potential inaccuracy of PR levels from IHC analysis may also be a contributing factor to this discrepancy.

Our study also hinted that the discordance between surrogate assay and genetic assays may be smaller than we expect. The discrepancy between intrinsic subtyping and surrogate assay is clearly recognized in the field. That is also the driven force for the campaign of universal genetic testing for breast cancer patients. However, in this study, by merely improving the accuracy of Ki67 measurement in surrogate assay, we have significantly improved the performance of surrogate assay. Future studies are urgently needed to compare various genetic assays including PAM50 with adjusted surrogate assay.

We are also optimistic that by measuring ER, Her2, and PR protein levels objectively and quantitatively, the performance of surrogate assay should be improved even further. In fact, we have demonstrated previously that using ER, PR and Her2 protein levels as absolutely and continuous variables, we were able to separate the clinical samples into three distinct groups closely matching the intrinsic subtypes (3D subtyping)(13). Future studies are needed to evaluate the prognostic performance of this 3D subtyping in daily clinical practice.

In summary, the Ki67 protein levels were measured unprecedentedly in 246 FFPE specimens absolutely, quantitatively and objectively. The measured Ki67 levels were used to replace the Ki67 scores from IHC analysis to adjust current prevailing surrogate assay using 2.31 nmole/g as cutoff, and we have demonstrated that the prognosis of adjusted surrogate assay was improved significantly for overall survival (OS) of Luminal-like patients. Thus, Ki67 levels are proposed to be measured using QDB method in daily clinical practice to improve the prognosis of surrogate assay.

## Data Availability

Data is available upon request by writing to

## Notes

### Competing Interest Statement

YLv, WZ, FT, YZ, JLv & JZ are employees of Yantai Quanticision Diagnostics, Inc., a division of Quanticision Diagnostics, Inc., who own or has filed patent applications for QDB plate, QDB method, & QDB application in IHC analysis. JH, JZou,SX, LJ, XW declared no conflict of interest

### Funding Statement

This research is funded by Yantai Quanticision Diagnostics, Inc.

